# A nationwide study of *Chlamydia trachomatis* infections in Denmark during the COVID-19 pandemic

**DOI:** 10.1101/2021.06.30.21259819

**Authors:** Paula L. Hedley, Steen Hoffmann, Ulrik Lausten-Thomsen, Marianne Voldstedlund, Karsten Dalsgaard Bjerre, Anders Hviid, Lone Krebs, Jørgen S. Jensen, Michael Christiansen

**Affiliations:** Department for Congenital Disorders, Statens Serum Institut, Copenhagen, Denmark; Department of Bacteria, Parasites & Fungi, Statens Serum Institut, Copenhagen, Denmark; Department of Neonatology, Copenhagen University Hospital Rigshospitalet, Copenhagen, Denmark; Data integration and Analysis, Statens Serum Institut, Copenhagen, Denmark; Department of Epidemiological Research, Statens Serum Institut, Copenhagen, Denmark; Department of Drug Development and Clinical Pharmacology, Pharmacovigilance Research Center, Faculty of Health and Medical Sciences, University of Copenhagen, Copenhagen, Denmark; Department of Gynecology and Obstetrics, Copenhagen University Hospital, Amager and Hvidovre Hospital, Copenhagen, Denmark; Department of Clinical Medicine, University of Copenhagen, Copenhagen, Denmark; Department of Biomedical Science, University of Copenhagen, Copenhagen, Denmark

**Keywords:** *Chlamydia trachomatis*, pregnancy infection, COVID-19, preterm birth

## Abstract

**Objectives:** COVID-19 policies have been employed in Denmark since March 2020. We examined whether COVID-19 restrictions had an impact on *Chlamydia trachomatis* infections compared with 2018 and 2019.

**Methods:** This retrospective nation-wide Danish observational study was performed using monthly incidences of laboratory confirmed chlamydia cases and number of tests, obtained from nation-wide surveillance data. Additionally, Oxford COVID-19 Government Response Tracker data, and Google COVID-19 Community Mobility Reports were used to contextualise the behavioural adaptions seen as a result of COVID-19 policies. Testing rates were compared using Poisson regression and test positivity rates were compared using logistic regression.

**Results:** The crude incidence rate (IR) of laboratory confirmed chlamydia infections was reduced to 66.5 per 10^5^ during the first (March-April 2020) lockdown period as compared to 88.3 per 10^5^ in March-April 2018-2019, but the testing rate was also reduced (Rate ratio 0.72 95% CI 0.71 – 0.73), whereas the odds ratio for a positive test between the two periods was 0.98 (95% CI 0.96 – 1.00). The period of eased COVID 19 restrictions (May – December 2020) and the second lockdown period (December 2020 – March 2021) were characterised by marginally increased crude IRs, while the number of tests performed, and test positivity rates returned very close to the levels seen in 2018-2019. These results were independent of sex, age group, and geographical location.

**Conclusion:** The first Danish COVID-19 lockdown resulted in a reduction in the number of chlamydia tests performed and a consequent reduction in the number of laboratory-identified cases. This period was followed by a return of testing and test positivity close to the level seen in 2018 – 2019. Altogether the Danish COVID-19 restrictions have had negligible effects on laboratory confirmed *C. trachomatis* transmission.

## Introduction

COVID-19 was declared a pandemic on March 11, 2020.^1^ In Denmark, a strict lockdown was implemented which restricted workplace and school attendance, while encouraging residents to stay home.^2^ COVID-19 restrictions have remained in place, with varying stringency ever since. These restrictions have had a plethora of behavioural and psycho-social effects.^3^

Danish COVID-19 restrictions have, as expected, been associated with remarkable reductions in the incidence of infectious diseases such as influenza, and pertussis, as well as invasive meningococcal and pneumococcal infections^4 5^. Perhaps less expected, COVID-19 restrictions have also been associated with a dramatic reduction in extremely preterm birth.^6^ This raises the question, could a reduction in infectious disease burden, particularly ascending urogenital infections (UGIs), several of which have been associated with preterm birth,^7^ explain the reduction in extremely preterm birth? While we can’t answer this question directly, we can assess the incidence of chlamydia within the Danish population as a proxy for transmissible UGIs.

In Denmark, chlamydia is the most frequent bacterial sexually transmitted infection (STI), and it has been subject to mandatory laboratory notification since 1994. Additionally, *Chlamydia trachomatis* infection either co-occurs or shares predisposing risk factors with several UGIs, e.g. bacterial vaginosis^8^ or genital mycoplasmas,^9^ which makes it a possible surrogate marker for a broader range of, non-notifiable, UGIs.

We hypothesized, that the incidence of *C. trachomatis* infections would reflect behavioural changes induced by the COVID-19 restrictions in Denmark. Using publicly available data originating from The Danish Microbiology Database (MiBa)^10^ via the webportal at Statens Serum Institut^11^ alongside national surveillance data pertaining to the number of tests performed,^12^ we compared the incidence of laboratory verified *C. trachomatis* infections since the initiation of COVID-19 restrictions with that of the preceding two years, 2018-2019. The incidences were contextualized using lockdown stringency information from the Oxford COVID-19 Government Response Tracker (OxCGRT)^13 14^ and mobility data from Google COVID-19 Community Mobility Reports.^15^

## Methods

### Data sources

Numbers of laboratory confirmed *C. trachomatis* infections were obtained via extracts from MiBa via the webportal at Statens Serum Institut.^10 11^ The monthly number of *C*.*trachomatis* laboratory tests performed since January 2018 were obtained from national surveillance data extracted from MiBa^12^. Lockdown stringency data were obtained from OxCGRT.^13 16^ Mobile phone-based location data were obtained from Google COVID-19 Community Mobility Reports.^15^ With the exception of the national surveillance data,^12^ all data were obtained from publicly available sources.

### Study periods

We have examined the period since the first containment policies were enacted following the report of the first Danish COVID-19 cases on February 27, 2020, until March 31, 2021 (Figure 1). The first wave lockdown (March 12 – April 14, 2020), was followed by a period where restrictions were eased just to be tightened again with increasing infection rate, second wave lockdown (December 17 – March 31, 2021) (Figure 1). The period of COVID-19 restrictions was compared with the aggregate data from the same calendar periods for the years 2018-2019. Gender and age distributions were compared with data from 2015-2019.

**Figure 1.**
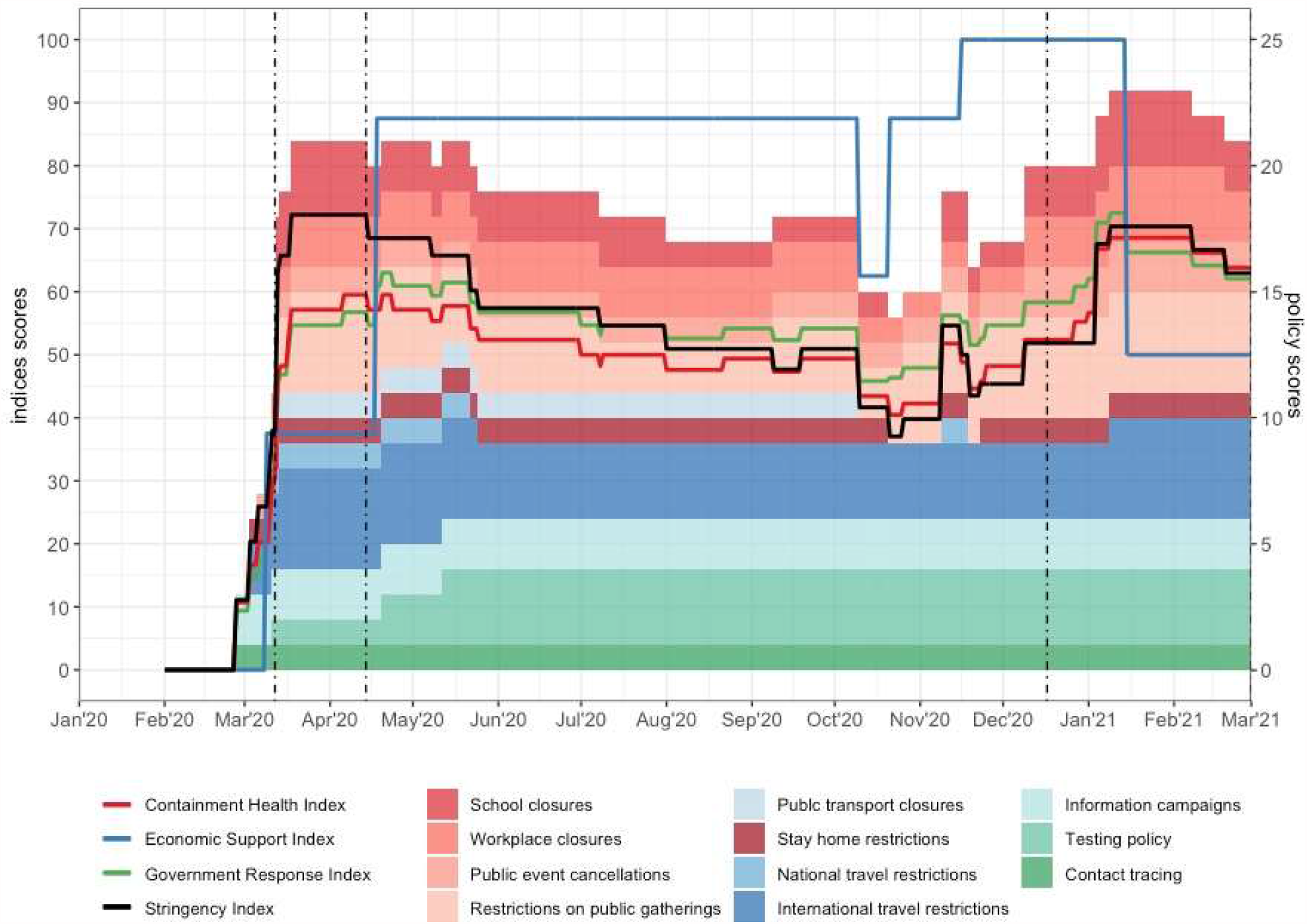
The Danish COVID-19 restrictions, the black line represents the overall stringency index, the stacked coloured columns represent the measures of the containment and control policies implemented during the whole COVID-19 restriction period. The coloured lines represent Containment, Economic support, and Government response indices, respectively. Data source for panel B: https://www.bsg.ox.ac.uk/research/research-projects/covid-19-government-response-tracker#data.^13^ The black dot-dashed vertical lines define the extent of the two strict lockdown periods.

### Statistics

We describe incidence rates (number of laboratory-confirmed *C. trachomatis* infections / population at risk on January 1st of the relevant year) according to age, sex, period, and geographical region (province of residence). The number of tests performed is described per month since January 2018. We compare the number of tests performed using Poisson regression, offset by population size to estimate rate ratios (RRs) with 95% confidence intervals (CIs). RRs were adjusted for period (2018-2019, 2020, and 2021). We estimated odds ratios (ORs) with 95% CIs comparing the odds of a positive test result (among all tests) according to period.

Statistics were conducted using R version 4.0.3.

### Ethics

All data were aggregate and most from publicly available sources supplemented with national surveillance data, so approvals from neither scientific ethics committees nor Data Authorities were necessary as per Danish law and regulations.

## Results

The crude incidence rate (IR) of laboratory confirmed chlamydia infection was 66.5 per 10^5^ during the first lockdown March-April 2020, as compared to a crude IR of 88.3 per 10^5^ in March-April 2018-2019 (Table 1). However, the adjusted Rate Ratio (aRR) of tests performed in the periods was 0.72 (95% CI 0.71 – 0.73), whereas the OR of having a positive test was 0.98 (95% CI 0.96 – 1.00) (Table 1). This, and the monthly number of tests performed (Supplementary Table 1) suggests that the reduced crude IR in the first lockdown period is caused by reduced testing. During the May-December 2020 restriction period and the second lockdown January 2020 – March 2021, the crude IRs of laboratory confirmed chlamydia infection were similar to the crude IRs of the 2018 – 2019 periods (Table 1). In the same periods the overall aRR of tests performed and the test positivity were similar to the values noted for 2018-2019 (Table 1 & Supplementary Table 1).

**Table 1:**
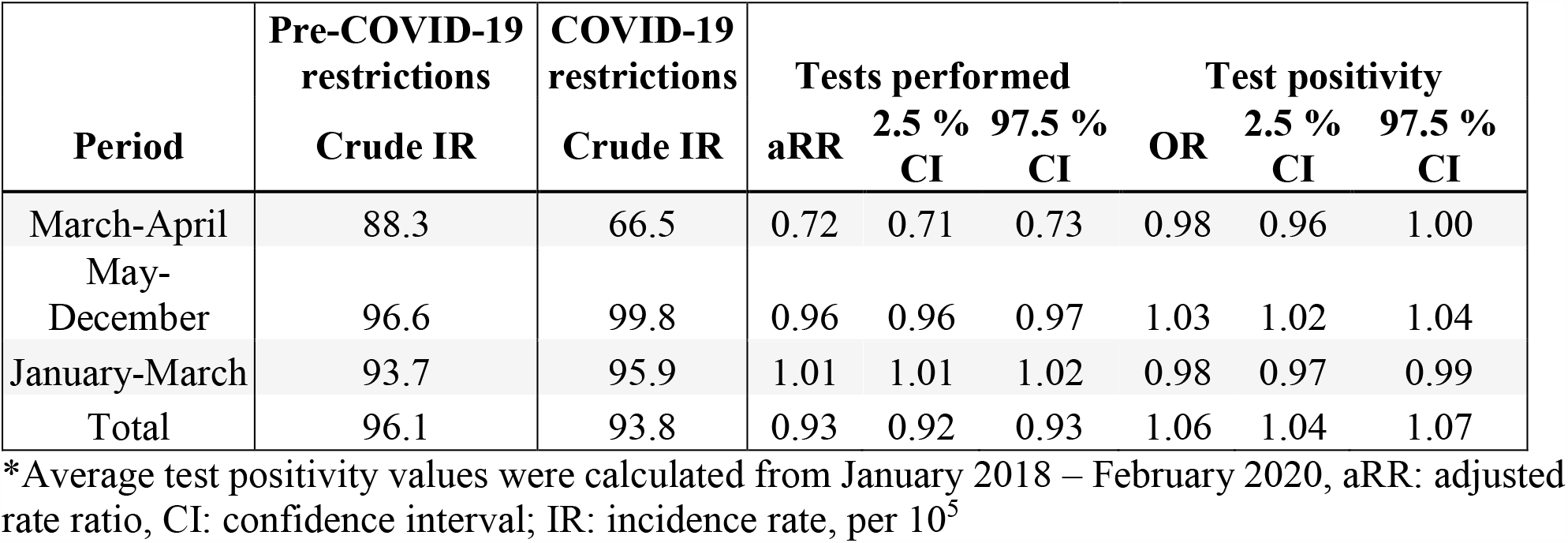
Incidence rates per COVID-19 restriction periods compared to the same periods in 2018-2019.

| Period | Pre-COVID-19<br>restrictions<br>Crude IR | COVID-19<br>restrictions<br>Crude IR | Tests performed |  |  | Test positivity |  |  |
| --- | --- | --- | --- | --- | --- | --- | --- | --- |
|  |  |  | aRR | 2.5 %<br>CI | 97.5 %<br>CI | OR | 2.5 %<br>CI | 97.5 %<br>CI |
| March-April | 88.3 | 66.5 | 0.72 | 0.71 | 0.73 | 0.98 | 0.96 | 1.00 |
| May-December | 96.6 | 99.8 | 0.96 | 0.96 | 0.97 | 1.03 | 1.02 | 1.04 |
| January-March | 93.7 | 95.9 | 1.01 | 1.01 | 1.02 | 0.98 | 0.97 | 0.99 |
| Total | 96.1 | 93.8 | 0.93 | 0.92 | 0.93 | 1.06 | 1.04 | 1.07 |

\*Average test positivity values were calculated from January 2018 – February 2020, aRR: adjusted rate ratio, CI: confidence interval; IR: incidence rate, per 10<sup>5</sup>

The Danish COVID-19 policies are indicated in Figure 1. The first Danish COVID-19 case was found on February 27, 2020, and this was followed by information campaigns, introduction of testing and contact tracing, travel restrictions, and other containment strategies^17^ (Figure 1). March 12, 2020, when the number of COVID-19 cases rose dramatically and the WHO declared COVID-19 a pandemic, strict lockdown measures were introduced, with workplace and school closures, as well as stay home restrictions. This lockdown period lasted until April 14, where a gradual easing commenced. This period of lessened COVID-19 restrictions, lasted until late December 2020, where the second wave of the pandemic necessitated a second lockdown that was still in effect at the end of March 2021.

The mobility information from Google (Supplementary Figure 1) demonstrates the behavioural consequences of the lockdown period, with a reduction (∼60 %) in workplace presence and an increase (∼18 %) in the proportion of time spent in places of residence during the first lockdown period. The second lockdown did not result in an effect similar to the first, as a reduction of only ∼30 % (except for a short reduction to ∼60 % during Christmas holidays) in workplace presence, and an increase of ∼12 % of time spend in places of residence during the second lockdown period were noted (Supplementary Figure 1). This behavioural difference was seen despite a similar level of formal lockdown stringency (Figure 1). Thus, the first lockdown period was associated with a reduction in testing for *C. trachomatis* as well as a reduction in laboratory confirmed chlamydia infections, followed by a relative increase in testing and number of infections between May and December 2020, whereas the second lockdown period was characterised by a marginal increase in the number of tests performed, and a marginal reduction in test positivity (Table 1 & Supplementary Table 1).

The monthly chlamydia incidence in women and men (Supplementary Figure 2) distributed in age groups, demonstrate, as seen in Supplementary Table 1, a reduction in laboratory confirmed cases during the first lockdown and an increase in July 2020. No significant deviation from the 2015-2019 numbers was seen in either sex or age-groups during the second lockdown from January 2020 – March 2021.

The chlamydia incidence in the 11 provinces of Denmark is shown in Supplementary Figure 3. No marked differences in temporal development between provinces were observed, suggesting that the behaviour changes associated with COVID-19 restrictions were uniformly spread throughout Denmark.

The ratio between the number of men and women with chlamydia was < 0.6 in persons aged 15 – 24 years but increased with age (Figure 2) to > 1 in persons aged 45 – 54 years. This picture is compatible with the pre-COVID lockdown epidemiology of chlamydia.^18^ However, a decrease in the ratio in March, followed by an increase in April and May 2020, in persons in the age group 45-54 years was noted.

**Figure 2.**
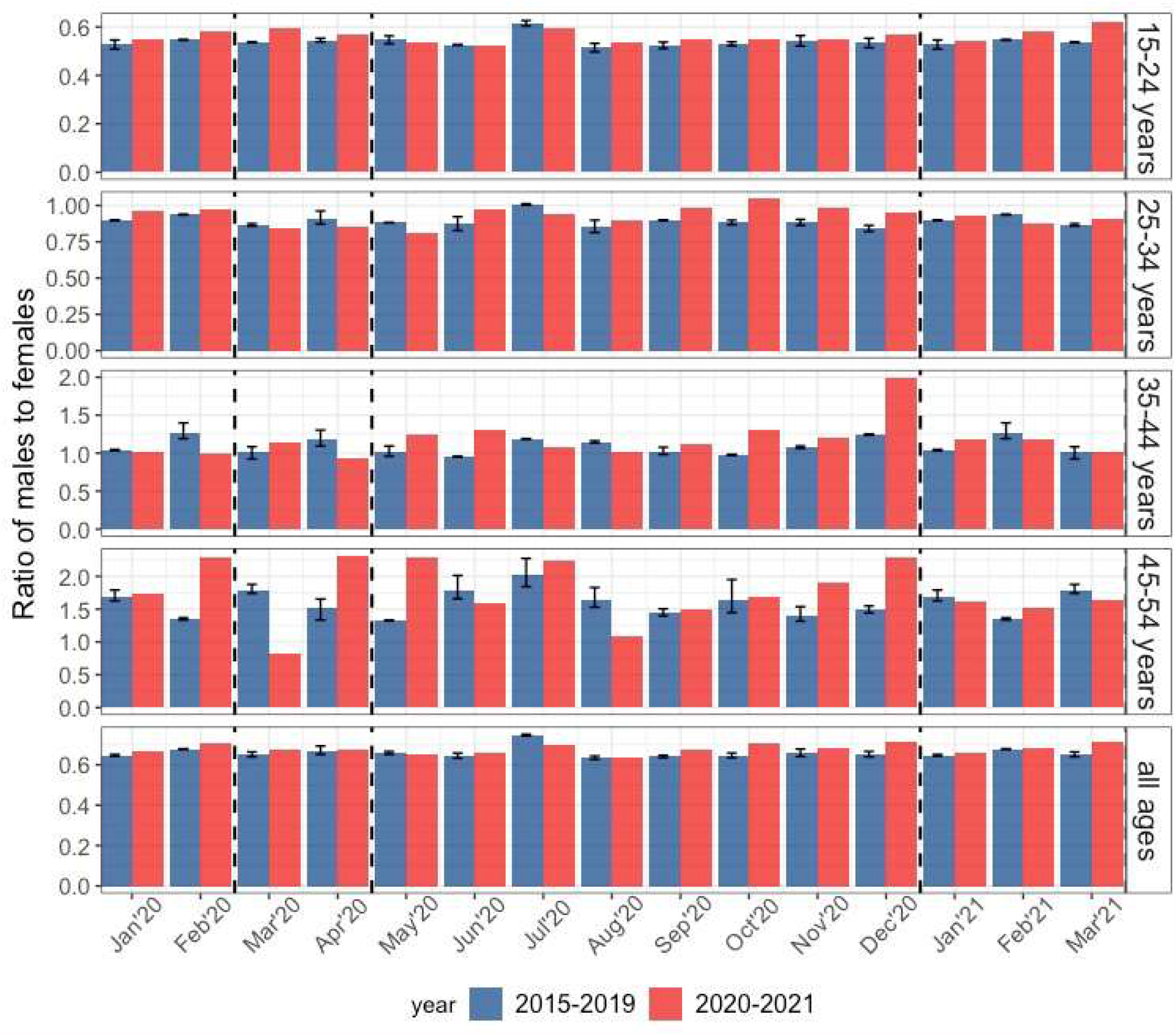
The ratio of males to females among the laboratory verified *Chlamydia trachomatis* infections in different age groups. Bars indicate upper 95%-confidence interval.

## Discussion

The first lockdown period was associated with a moderate, ∼25 %, decrease in laboratory confirmed *C. trachomatis* cases (Table 1). However, this period was also associated with a similar decrease in laboratory tests performed (Table 1) and accounting for that showed that in fact there was no change in the odds of receiving a laboratory confirmed chlamydia diagnosis during the first lockdown and the average odds for the same period between 2018 and 2019 (Table 1). This suggests that the decrease in incidence rate is caused by reduced testing. The second lockdown showed a nominal increase in chlamydia cases (Table 1) despite being formally as stringent as the first lockdown (Figure 1). However, the number of tests performed also increased marginally during the second lockdown and after accounting for this there is a small reduction between the odds of receiving a laboratory confirmed chlamydia diagnosis during the second lockdown and the corresponding odds during the same period in 2018-2019. This may suggest that the behavioural response was markedly more relaxed during the second lockdown than during the first lockdown (Supplementary Figure 1).

The period between the lockdowns was associated with a marginal increase in the incidence rate of laboratory confirmed chlamydia driven by an increase in test positivity between June and September, possibly explained by delayed testing. Indeed, in mid-March the Danish Organization of General Practitioners and the Danish Society of General Medicine issued a recommendation,^19^ that only patients with acute disease should attend the GP’s surgery, and consultations should preferentially be conducted over the phone. Which may have resulted in some delay in testing, particularly as 70-80 % of chlamydia infections in Denmark are asymptomatic, and most tests are performed as part of opportunistic screening. The restrictions for attending GPs were lifted in April, making it worthwhile considering whether planned or delayed tests were simply done over the following months. Additionally, the statement that “*Sex is good, sex is healthy, the Danish Health Authority is in favour of sex*”, purported broadly by the director of the Danish Health Authority in the Danish media in April 2020 ^20^ may have contributed to a normalisation of sexual behaviours and consequent normalisation of chlamydia cases. It cannot be excluded that the decrease in testing, and consequent decrease in incidence of laboratory confirmed cases, seen in March - April 2020 was caused - at least partly - by reduced sexual activity among teens and young adults during the first lockdown; however, the period also saw a marked increase in online dating internationally,^21^ as well as in Denmark.^22^, suggesting that reduced sexual activity was not the explanation. The gender distribution of laboratory-verified cases of chlamydia during the COVID-19 restrictions was similar to that seen the years previously, suggesting that the overall epidemiology was unchanged (Figure 2). Likewise, the temporal variation in chlamydia incidence during the COVID-19 restrictions was neither influenced by gender (Supplementary Figure 2), nor geographical location (Supplemental Figure 3).

Overall, the Danish COVID-19 restrictions – during the first lockdown – were associated with a decrease in the extent of testing and, consequently, of incidence of laboratory confirmed chlamydia. The COVID-19 restrictions had very little – if any – effect on transmission of chlamydia in Denmark. Similar findings were reported from Finland, where *C. trachomatis* and *Neisseria gonorrhoeae* incidence were completely unperturbed during the first five months of the pandemic and lockdown measures.^23^ This is in contrast to cities in Italy and Spain, where the number of STIs (both syphilis, gonorrhoea and chlamydia) dropped precipitously during the lockdown.^24 25^ However, in Trento, Italy, the number of cases in 2020 was comparable to 2019, and risky sexual behaviour was not reduced.^26^ The difference between parts of Italy and Spain and northern Europe may be the extent to which tourism contributes to the incidence of STIs,^24^ as well as the extent to which the population follows the recommendations and instructions by the health authorities. In the US, a slightly different pattern was seen, with a decrease in chlamydia of ∼20 % compared to 2019, and a smaller decrease in gonorrhoea and an unchanged syphilis incidence.^27^ Perhaps the significant reductions in all STIs depend on whether a curfew is part of the lockdown restrictions? The lack of relation between limitations in mobility and social interaction and chlamydia incidence may be a result of the extent to which dating apps were used, particularly important among young adults.^21 28^ In conclusion, the first Danish lockdown period was associated with a decrease in testing and a subsequent reduced incidence of laboratory confirmed chlamydia. This was not seen in the second lockdown period and the intervening period of eased restrictions. Thus, the period of COVID-19 restrictions, with its significant effects on behaviour, was not associated with appreciable changes in chlamydia transmission.

## Data Availability

Data on incidence of positive chlamydia tests are publically available from the MiBa SSI webportal (www.ssi.dk). Data on test number were obtained from data not yet processed for the MiBa database.

## Acknowledgements

The research was conducted using the resources of the Danish National Biobank, funded by the Novo Nordisk Foundation.

## Legends to figures and supplemental figures

**Supplementary Figure 1.**
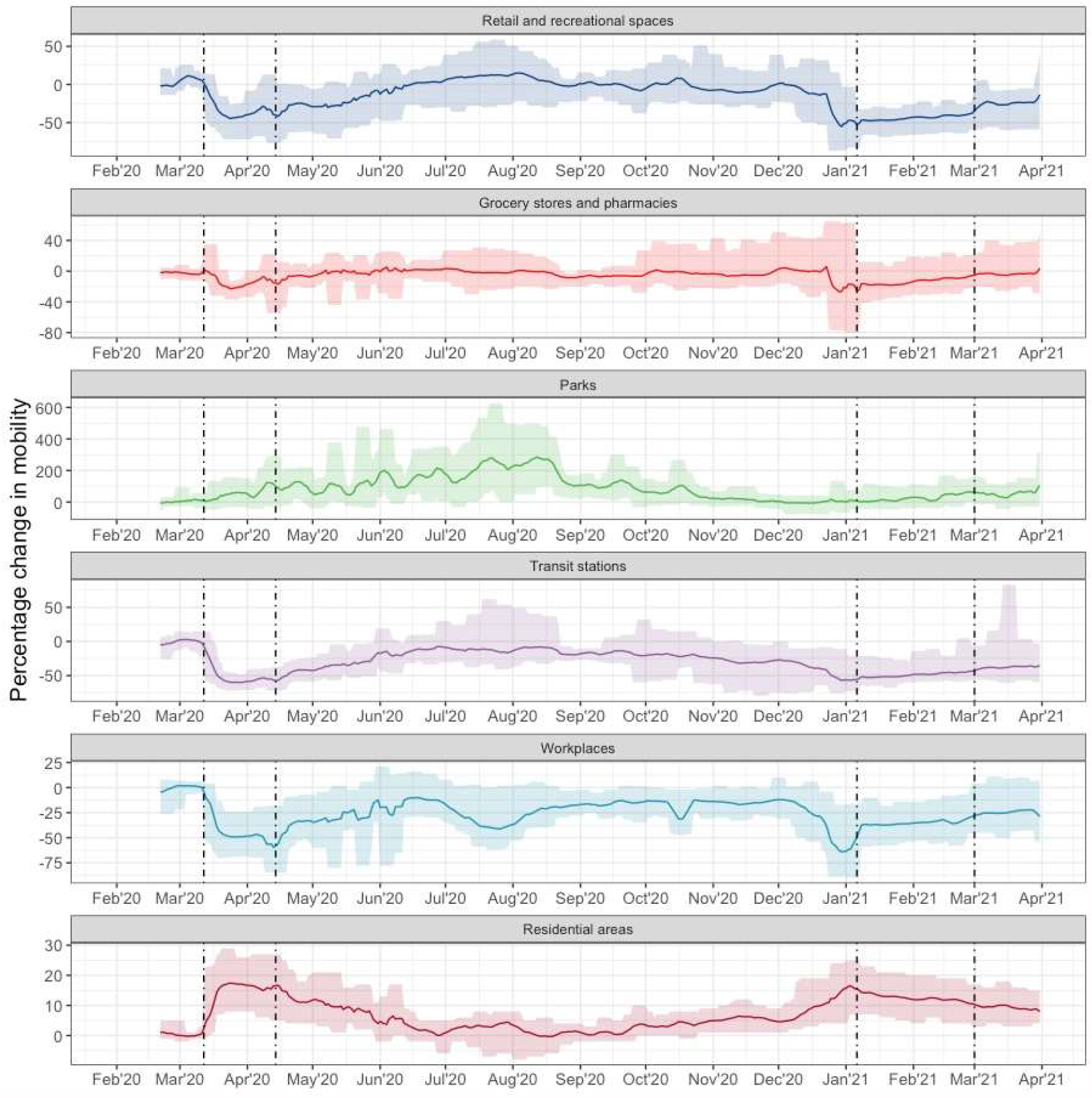
The daily average mobile-phone based mobility of residents of Denmark. Percentage changes in mobility are presented with reference to a baseline location assessment (the median values from the 5-week period January 3 – February 6, 2020). Changes in total visits to workplaces (light blue), retail and recreational spaces (dark blue), grocery stores and pharmacies (light red), parks (green), and Transit stations (grey), as well as time spent in places of residence (dark red). The black vertical lines define the extent of the two strict lockdown periods. Data source: Google Mobility Reports.^15^

**Supplementary Figure 2.**
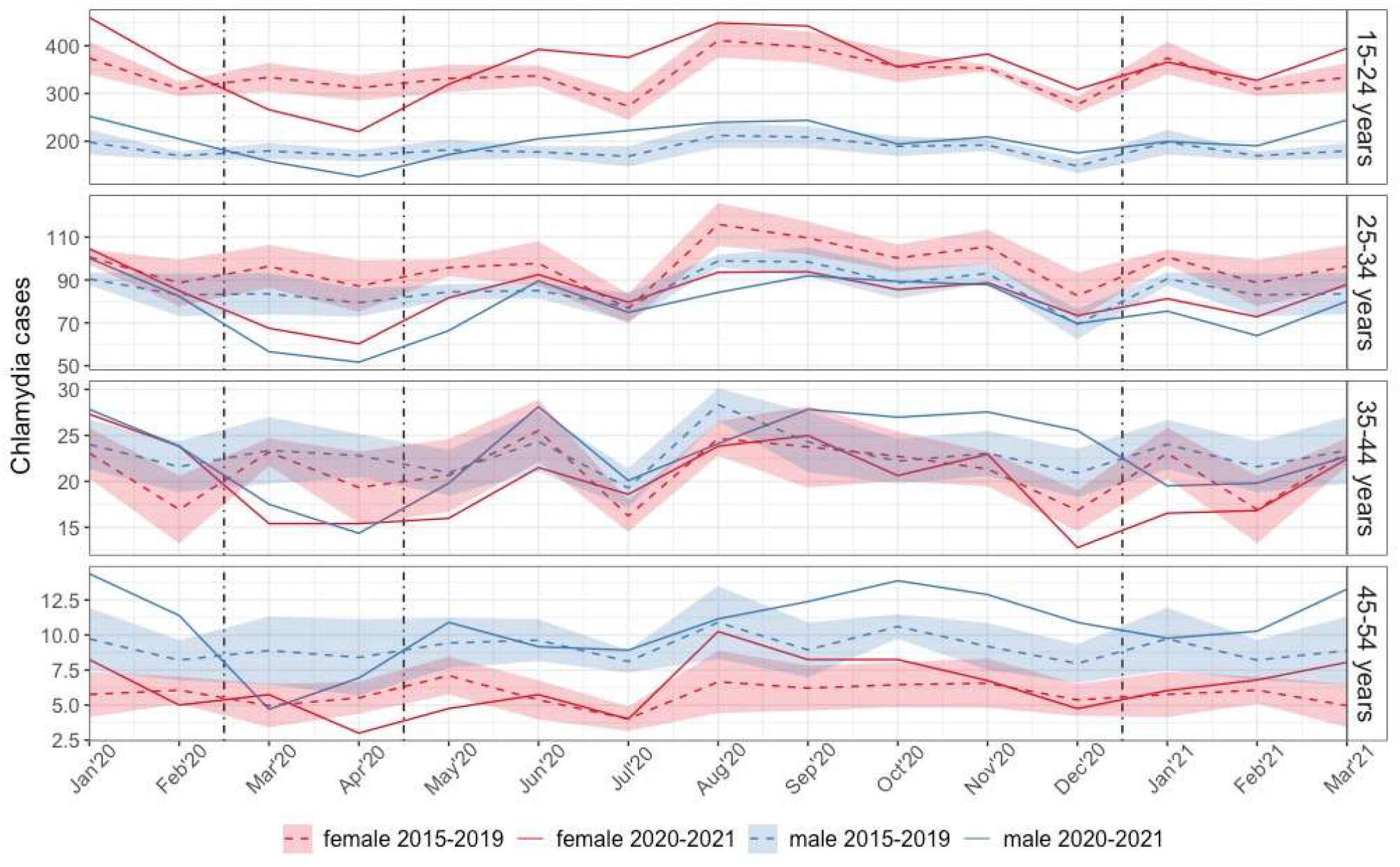
Monthly incidence of Chlamydia cases in **A**. males and **B**. females in specific age groups. The red line in the monthly incidence in 2020-21, and the blue line and shade is the average incidence from 2015-2019 +/-1 SD. The black vertical lines define the extent of the two strict lockdown periods

**Supplementary Figure 3.**
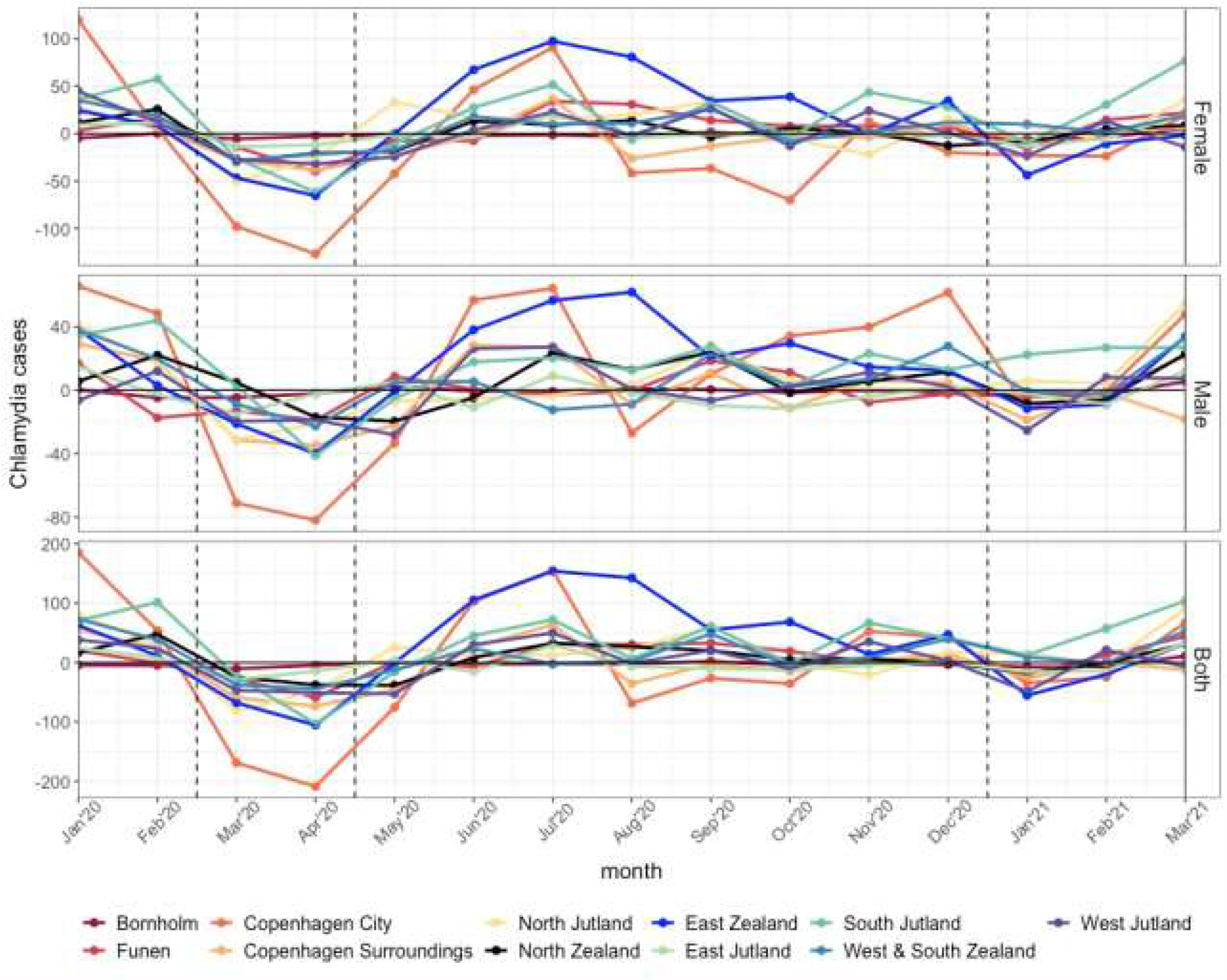
The monthly incidence of *Chlamydia trachomatis* cases in the 11 provinces of Denmark. The data are divided into males and females, and the total incidence is shown in the bottom panel. The black vertical lines define the extent of the two strict lockdown periods.

## Supplementary Materials

**Supplementary Table 1:**
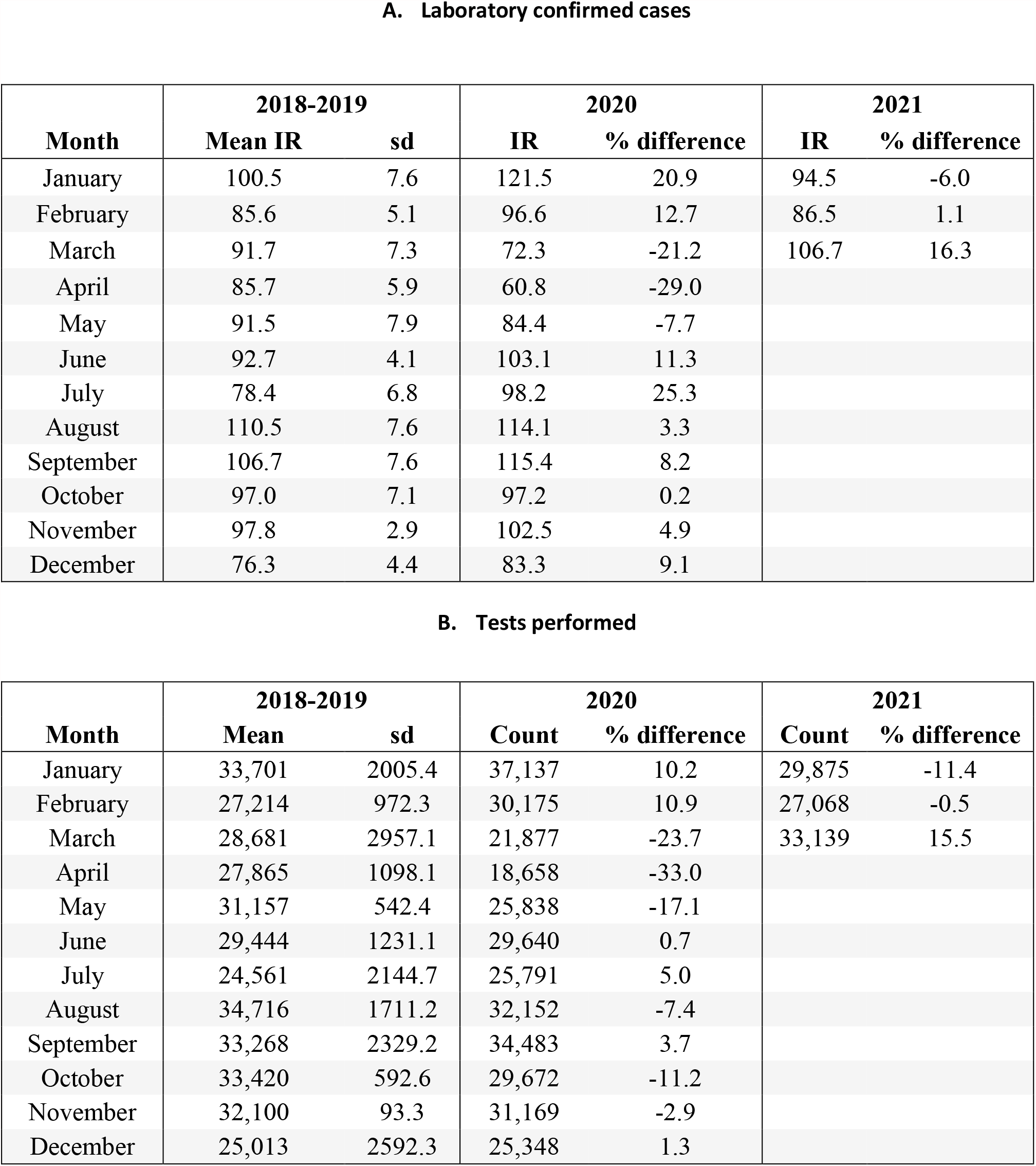
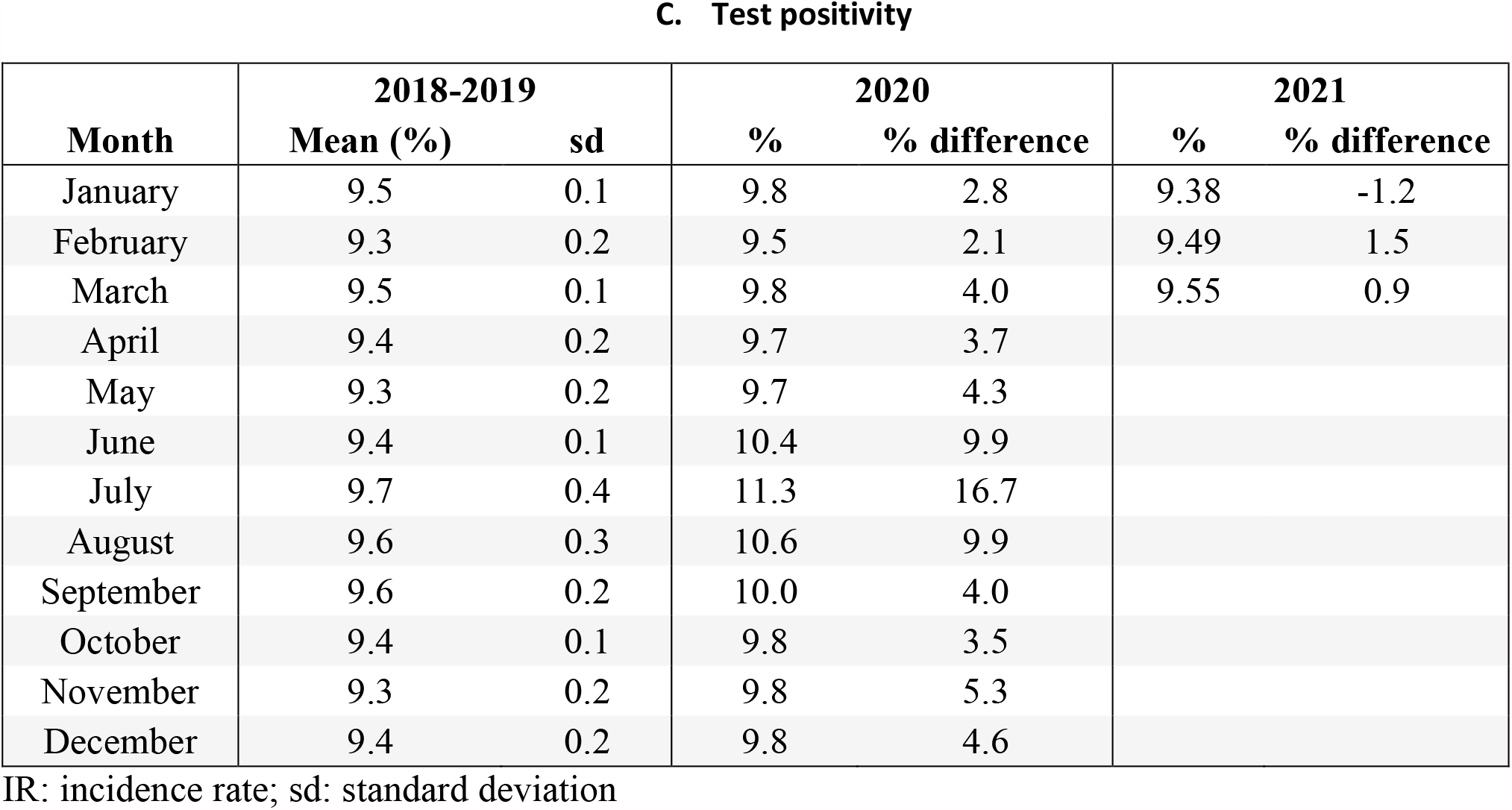
**Description of A. the monthly laboratory confirmed chlamydia case incidence rate (IR) (per 10**^**5**^**), B. number of tests performed, and C. test positivity rates. % difference shows the difference between year of interest and 2018-2019 for each month**.

A. Laboratory confirmed cases
| Month | 2018-2019 |  | 2020 |  | 2021 |  |
| --- | --- | --- | --- | --- | --- | --- |
|  | Mean IR | sd | IR | % difference | IR | % difference |
| January | 100.5 | 7.6 | 121.5 | 20.9 | 94.5 | -6.0 |
| February | 85.6 | 5.1 | 96.6 | 12.7 | 86.5 | 1.1 |
| March | 91.7 | 7.3 | 72.3 | -21.2 | 106.7 | 16.3 |
| April | 85.7 | 5.9 | 60.8 | -29.0 |  |  |
| May | 91.5 | 7.9 | 84.4 | -7.7 |  |  |
| June | 92.7 | 4.1 | 103.1 | 11.3 |  |  |
| July | 78.4 | 6.8 | 98.2 | 25.3 |  |  |
| August | 110.5 | 7.6 | 114.1 | 3.3 |  |  |
| September | 106.7 | 7.6 | 115.4 | 8.2 |  |  |
| October | 97.0 | 7.1 | 97.2 | 0.2 |  |  |
| November | 97.8 | 2.9 | 102.5 | 4.9 |  |  |
| December | 76.3 | 4.4 | 83.3 | 9.1 |  |  |

B. Tests performed
| Month | 2018-2019 |  | 2020 |  | 2021 |  |
| --- | --- | --- | --- | --- | --- | --- |
|  | Mean | sd | Count | % difference | Count | % difference |
| January | 33,701 | 2005.4 | 37,137 | 10.2 | 29,875 | -11.4 |
| February | 27,214 | 972.3 | 30,175 | 10.9 | 27,068 | -0.5 |
| March | 28,681 | 2957.1 | 21,877 | -23.7 | 33,139 | 15.5 |
| April | 27,865 | 1098.1 | 18,658 | -33.0 |  |  |
| May | 31,157 | 542.4 | 25,838 | -17.1 |  |  |
| June | 29,444 | 1231.1 | 29,640 | 0.7 |  |  |
| July | 24,561 | 2144.7 | 25,791 | 5.0 |  |  |
| August | 34,716 | 1711.2 | 32,152 | -7.4 |  |  |
| September | 33,268 | 2329.2 | 34,483 | 3.7 |  |  |
| October | 33,420 | 592.6 | 29,672 | -11.2 |  |  |
| November | 32,100 | 93.3 | 31,169 | -2.9 |  |  |
| December | 25,013 | 2592.3 | 25,348 | 1.3 |  |  |

**C. Test positivity**
| **Month** | **2018-2019** |  | **2020** |  | **2021** |  |
| --- | --- | --- | --- | --- | --- | --- |
|  | **Mean (%)** | **sd** | **%** | **% difference** | **%** | **% difference** |
| January | 9.5 | 0.1 | 9.8 | 2.8 | 9.38 | -1.2 |
| February | 9.3 | 0.2 | 9.5 | 2.1 | 9.49 | 1.5 |
| March | 9.5 | 0.1 | 9.8 | 4.0 | 9.55 | 0.9 |
| April | 9.4 | 0.2 | 9.7 | 3.7 |  |  |
| May | 9.3 | 0.2 | 9.7 | 4.3 |  |  |
| June | 9.4 | 0.1 | 10.4 | 9.9 |  |  |
| July | 9.7 | 0.4 | 11.3 | 16.7 |  |  |
| August | 9.6 | 0.3 | 10.6 | 9.9 |  |  |
| September | 9.6 | 0.2 | 10.0 | 4.0 |  |  |
| October | 9.4 | 0.1 | 9.8 | 3.5 |  |  |
| November | 9.3 | 0.2 | 9.8 | 5.3 |  |  |
| December | 9.4 | 0.2 | 9.8 | 4.6 |  |  |

IR: incidence rate; sd: standard deviation

